# Within-hotel transmission of SARS-CoV-2 during on-arrival quarantine in Hong Kong

**DOI:** 10.1101/2022.06.25.22276894

**Authors:** Dillon C. Adam, Mario Martín-Sánchez, Haogao Gu, Peng Wu, Eric H. Y. Lau, Gabriel M. Leung, Leo L. M. Poon, Benjamin J. Cowling

## Abstract

**Background:** On-arrival quarantine has been one of the primary measures used to prevent the introduction of COVID-19 into Hong Kong since the start of the pandemic. Most on-arrival quarantines have been done in hotels, with the duration of quarantine and testing frequency during quarantine varying throughout the pandemic for various reasons. However, hotels are not necessarily designed with infection control in mind. We aimed to study the potential risk of transmission between persons in on-arrival quarantine.

**Methods:** We examined data on each laboratory-confirmed COVID-19 case identified in on-arrival quarantine in a hotel in Hong Kong between 1 May 2020 and 31 January 2022. We sequenced the full genomes of viruses from cases that overlapped with other confirmed cases in terms of the hotel of stay, date of arrival and date of testing positive. A combination of epidemiological information and sequence information was then used to identify probable transmission events.

**Findings:** Among 221 imported cases that overlapped with other quarantined cases, phylogenetic analysis identified eight suspected clusters comprising 20 cases in total. Only three of these clusters had been recognised as hotel transmission events.

**Interpretation:** We have identified potential occurrences of COVID-19 transmission within hotel quarantine in Hong Kong demonstrating the underlying low but non-zero risk associated with sequestering arrivals within hotels. In future pandemics, on-arrival quarantine in hotels could be used to delay the introduction of infection, but the construction of purpose-built facilities for on-arrival quarantine might be necessary to minimize importation risk.

**Funding:** Health and Medical Research Fund, Hong Kong

## INTRODUCTION

One of the measures used to reduce importation of COVID-19 into a locality is quarantine of arriving persons. In Hong Kong, quarantine of arrivals either at home, within purpose-built quarantine facilities or hotel quarantine (HQ) has been variably implemented since 27 January 2020, with mandatory quarantine for all arrivals since 19 March 2020. Quarantine within hotels for 14 days became mandatory for arrivals from some locations on 25 July 2020 and for all arrivals from 13 November 2020 with few exceptions. From 22 December 2020 onwards the Hong Kong government issued a list of selected hotels that were designated to receive inbound travellers, termed “designated hotel quarantine” (DHQ). Stringent infection control measures were implemented in these designated hotels.^1,2^ The required quarantine period increased to 21 days from 25 December 2020 as part of the responses to the emergence of SARS-CoV-2 variants.

Together with a range of local control measures, importation control has resulted in relatively limited local circulation of SARS-CoV-2 in Hong Kong in the first two years of the pandemic. By 31 January 2022, there had been approximately 13,000 confirmed cases, corresponding to just 1.9 confirmed cases per 1000 population in four distinct epidemic waves.^3,4^ However, in January 2022, spread of Omicron BA.2 occurred in Hong Kong and was not controlled, resulting in a large fifth epidemic wave with more than 1.1 million confirmed cases and more than 9000 deaths. ^5^ A small number of infections have occasionally been traced back to infections between guests or staff working within HQ/DHQ in Hong Kong.^6,7^ Transmission of infection from quarantined guests to other guests or staff has also been reported elsewhere in the world.^8,9^ Regular testing of hotel guests and staff can help to identify infections more quickly and limit leaks into the community, but it can be difficult to identify transmission between quarantined persons because those who test positive in HQ/DHQ would typically be assumed to have acquired their infection before they entered quarantine. In this study we combined genomic and epidemiological data to investigate the risk of infection within HQ/DHQ from 1 May 2020 through to 31 January 2022.

## METHODS

### Data collection and background

Line lists including demographic data were provided by the Centre for Health Protection (CHP) of the Department of Health for all cases confirmed by real time-polymerase chain reaction (RT-PCR) in Hong Kong from 1 May 2020 through to 31 January 2022. Cases were characterised as “imported” if they had a travel history overseas during the 21-days prior to the diagnosis and were presumed to have acquired the infection outside of Hong Kong (i.e. with no reason to presume otherwise). Arrivals were classified into seven regions of origin according to World Bank Development indicators given the most recent country of departure.

During the study period, all persons arriving in Hong Kong were tested upon arrival and then during quarantine (if negative at arrival) by RT-PCR. During the HQ period (up to and including 21 December 2020), guests were tested once before release towards the end of the 14-day quarantine period, whereas during the DHQ period (from 22 December 2020 onwards), guests were tested by PCR up to six times during quarantine, though testing frequency varied for many reasons including the length of quarantine, country of origin, and local testing constraints. During both periods, all laboratory-confirmed cases including asymptomatic cases and mild cases were isolated in hospital or in specialised isolation facilities. Based on the moment of detection and the place of stay after arrival, cases were classified into (a) detected at arrival within the place of specimen collection, (b) cases with home quarantine arrangements (e.g. cases coming from countries classified as low risk arriving before 13 November 2020), (c) cases within a specialised quarantine facility for high-risk arrivals (e.g. arrivals deemed close contacts of imported cases), (d) cases that stayed in HQ (either for quarantine purposes or while waiting for initial RT-PCR results in case of evening arrivals), and (e) cases under special quarantine arrangements (e.g. pilots and diplomats).

### Characterisation of potential HQ/DHQ clustering

For cases that stayed in HQ/DHQ, we obtained the period of stay using the date of arrival and the date of admission to isolation facilities. We considered cases for further investigation of SARS-CoV-2 infection or transmission during their quarantine period if they stayed at least one day in the same hotel as another case with overlapping periods of stay but not in the same room.

### Genomic sequencing of SARS-CoV-2

Saliva samples or nasopharyngeal swab samples of SARS-CoV-2 cases were sequenced. We reverse transcribed virus with primers targeting different regions^10^ with synthesized cDNA then subjected to multiple overlapping 2-kb PCRs for full-genome amplification. Where PCR amplicons were obtained from the same specimen, amplicons were pooled and sequenced using Nova sequencing platform (PE150, Illumina). The sequencing library was prepared by Nextera XT. Base calling of raw read signal and demultiplexing of reads by different samples were performed using Bcl2Fastq (Illumina). A reference-based re-sequencing strategy was applied in analysing the NGS data. Raw FASTQ reads were assembled and mapped to the SARS-CoV-2 reference genome (Wuhan-Hu-1, GenBank: MN908947.3) using BWA mem2 (v.2.0pre2). The primer reads and low-quality reads were trimmed using iVar^11^ with the above primers and default parameters. The consensus sequences for each sample were called as dominant bases at each position by samtools mpileup (v.1.11)^12^ and iVar with default parameters. Samples less than 27kb in length (excluding gaps) were excluded from further downstream analysis.

### Phylogenetic analysis & analysis of HQ/DHQ clustering

We selected 350 available genome sequences for analysis given the criteria of epidemiological overlap defined previously. We aligned genomes with greater than 70% unambiguous nucleotides (n=280) against A and B lineage references strains Wuhan/WH04/2020 and Wuhan-Hu-1/2019 respectively using MAFFT v7.49 for closely related viral genomes^10^ (FFT-NS-2 algorithm). Maximum likelihood phylogenetic trees were generated from the included alignment using IQ-Tree v2.1.2^13^ with 1000 ultrafast bootstrap replicates.^14^ We inferred sequence relatedness as the pairwise genetic distance between two sequences, calculating the number of genome-wide substitutions (HKY85 substitution model) given an alignment total length of 29,924 base pairs. Lineage assignment was determined using PANGOLIN v.3.1.14 and PLEARN model v.1.2.81 including designation of variants of concern (VOC).^15^

Probable transmission was determined between pairs identified by our initial screen based on epidemiological data if sequences were identical i.e., zero genome-wise substitutions, with the same country of departure, or ≤4 genome-wide substitutions between pairs with different countries of departure and were monophyletically exclusive. The substitution cut-off of ≤4 was determined based on the one-tailed 75% quantile of genetic distance between known close contacts also identified in HQ/DHQ (Supplementary Figure 1), with more strict criteria chosen to exclude potential false positives where sequences from the same country of departure were genomically similar yet unrelated to within HQ/DHQ transmission (e.g., in-flight or pre-arrival transmission). For all the above criteria, there must be epidemiological overlap given the hotel of stay, date of arrival, date of symptom onset and/or date of confirmation relative to each potential HQ/DHQ pair.

### Role of the funding source

Funders had no role in study design; in the collection, analysis, and interpretation of data; in the writing of the report; and in the decision to submit the paper for publication. Ethical approval was obtained from the Institutional Review Board of the University of Hong Kong.

## RESULTS

### Epidemiology of imported SARS-CoV-2 cases & sequence availability

During the study period, 13,165 cases were confirmed in Hong Kong, of which 23.9% (n=3152/13165) cases were classified as imported, i.e. presumed to have acquired infection whilst overseas, while the remaining 76.1% (n=10013/13165) of cases were determined to have acquired COVID-19 infection locally. Of the 3,152 SARS-CoV-2 cases confirmed in Hong Kong but classified as imported during the study period, the majority were aged between 20-39 years (51.9%, n=1634/3150, overall median = 34). Most imported cases arrived from South Asia (32.7%, n=1029/3150) followed by East Asia & the Pacific (30.1%, n=949/3150) and Europe & Central Asia (21.7%, n=685/3150) (Table 1).

**Table 1.** Descriptive epidemiology of SARS-CoV-2 cases confirmed in Hong Kong and characterised as overseas acquired cases from 1 May 2020 – 31 January 2022

| Descriptive variables | HQ/DHQ with overlap |  | HQ /DHQ cases | Non- HQ /DHQ cases | Total Imported |
| --- | --- | --- | --- | --- | --- |
|  | Seqs (n=221) | Total (n=720) | Total (n=1,189) | Total (n=1,963) | (n=3,152) |
| *Sex* |  |  |  |  |  |
| Female | 128 (57.9%) | 445 (61.9%) | 659 (55.4%) | 985 (50.2%) | 1644 (52.2%) |
| Male | 93 (42.1%) | 275 (38.1%) | 530 (44.6%) | 978 (49.8%) | 1508 (47.8%) |
| *Age* |  |  |  |  |  |
| 0-19 | 31 (14.0%) | 87 (12.1%) | 151 (12.7%) | 255 (13.0%) | 406 (12.9%) |
| 20-39 | 92 (41.6%) | 376 (52.3%) | 598 (50.3%) | 1036 (52.8%) | 1634 (51.8%) |
| 40-59 | 78 (35.3%) | 200 (27.8%) | 354 (29.8%) | 546 (27.8%) | 900 (28.6%) |
| 60-79 | 19 (8.6%) | 52 (7.1%) | 80 (6.7%) | 121 (6.2%) | 201 (6.4%) |
| 80+ | 1 (0.5%) | 5 (0.7%) | 6 (0.5%) | 5 (0.3%) | 11 (0.3%) |
| *Import region* |  |  |  |  |  |
| East Asia & Pacific | 78 (35.3%) | 300 (41.6%) | 400 (33.6%) | 550 (28.0%) | 950 (30.1%) |
| Europe & Central Asia | 21 (9.5%) | 82 (11.4%) | 242 (20.4%) | 443 (22.6%) | 685 (21.7%) |
| Latin America & Caribbean | 0 (0.0%) | 2 (0.3%) | 7 (0.6%) | 26 (1.3%) | 33 (1.0%) |
| Middle East & North Africa | 4 (1.8%) | 20 (2.8%) | 46 (3.9%) | 71 (3.6%) | 117 (3.7%) |
| North America | 14 (6.3%) | 38 (5.3%) | 91 (7.7%) | 143 (7.3%) | 234 (7.5%) |
| South Asia | 97 (43.9%) | 257 (35.7%) | 357 (30.0%) | 672 (34.2%) | 1029 (32.7%) |
| Sub-Saharan Africa | 5 (2.3%) | 16 (2.2%) | 35 (2.9%) | 41 (2.1%) | 76 (2.4%) |
| Missing | 2 (0.9%) | 5 (0.7%) | 11 (0.9%) | 17 (0.9%) | 28 (0.8%) |

Confirmed SARS-CoV-2 arrivals averaged 150 cases per month (Supplementary Figure 1) and peaked at 521 cases during the month of January 2022 following the global emergence of the Omicron variant and was lowest during May 2020 (n=42). Most cases classified as overseas acquired (62.3%, n=1961/3150) did not stay in HQ/DHQ but rather tested positive upon arrival at the airport (83.1%, n=1630/1961) or after arrival (16.9%, n=331/1961) either waiting for arrival results, home quarantine arrangements or within a specialised quarantine facility for high-risk arrivals. The remaining 37.8% (n=1189/3150) of cases classified as overseas acquired passed on-arrival testing and stayed within HQ/DHQ. We identified 60.6% (n=720/1189) of those cases having epidemiological overlap with other cases (Supplementary Figure 1). There was evidence that time from arrival-to-case detection among HQ/DHQ cases identified with epidemiological overlap (but not verified) was longer compared to those without overlap (median = 9 days vs. 4 days respectively, p<0.001, Wilcoxon rank sum test with continuity correction).

### Suspected transmissions within HQ/DHQ

Of the 720 overlapping cases detected in HQ/DHQ that we identified for further genomic analysis, we identified 147 potential clusters. Valid genomic samples (>70% unambiguous nucleotides) however were only available for 38.9% (n=280/720) of those cases corresponding to 95 (64.6%, n=95/147) possible clusters, though only 49 of those clusters comprising 221 cases could be analysed as the remaining clusters had only one valid genome sample or comprised only samples of known family contacts travelling together.

Given our previous genomic criteria, our analysis initially identified 13 initial clusters of within HQ/DHQ transmission. Further investigation found five of these clusters comprised sequences with identical countries of departure and date of arrival in Hong Kong thus indicating infection prior to HQ/DHQ could not be excluded, resulting in eight suspected HQ/DHQ clusters (Fig.1) comprising 20 cases.

**Figure 1.**
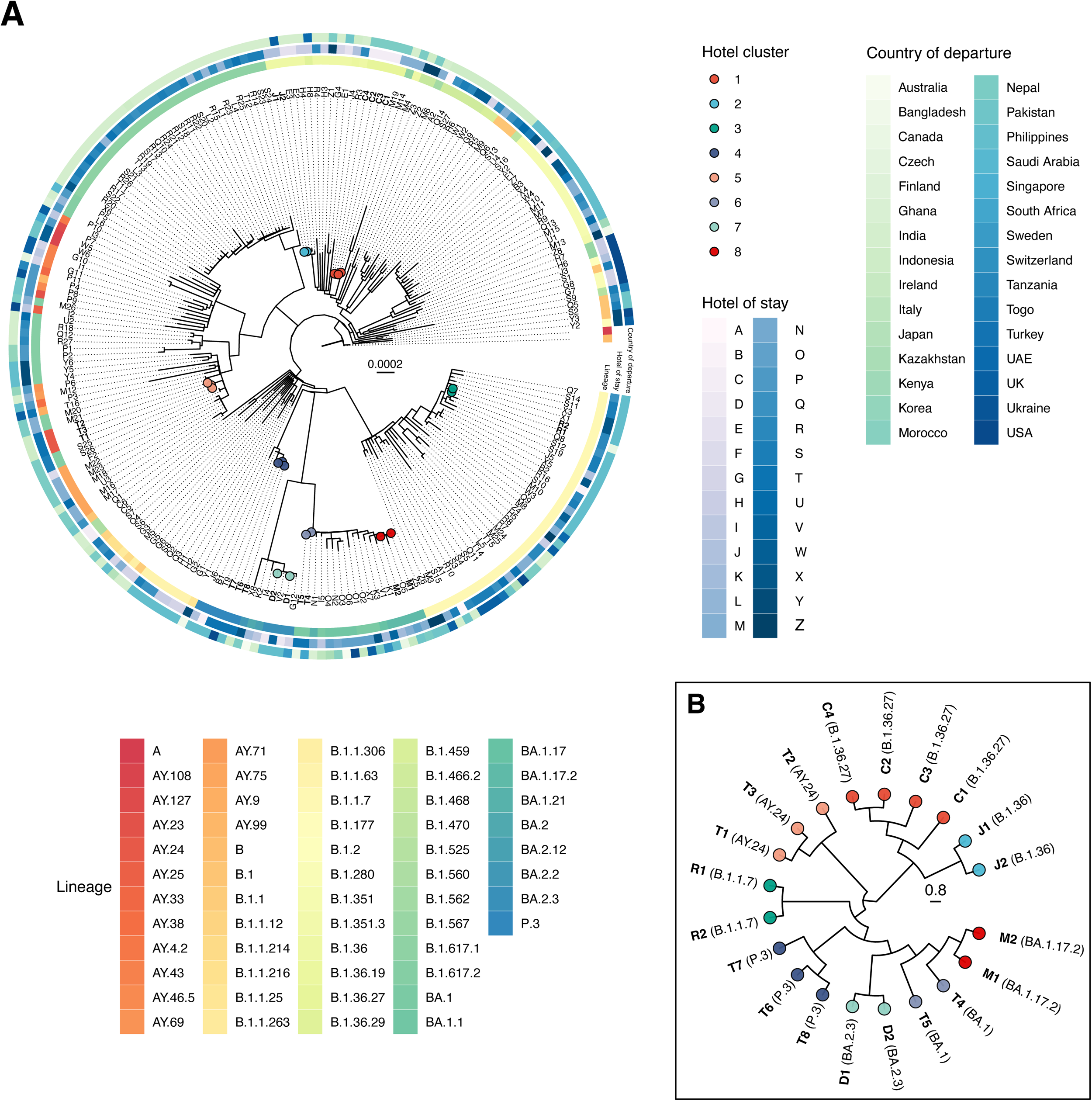
**A)** Phylogenetic tree of (n=221) all available cases of SARS-CoV-2 confirmed within hotel quarantine (HQ) or designated hotel quarantine (DHQ) with overlap to other unlinked cases in Hong Kong from 1 May 2020 through 31 Jan 2022 aligned against representative sequences of SARS-CoV-2 lineages A and B. Tips are coloured as confirmed within DHQ transmission clusters and labelled with unique case identifiers equivalent to hotel of stay. Radial heat map shows PANGO lineage, hotel of stay, and country of departure. **B)** Cladogram of suspected HQ/DHQ clusters.

Cluster one (contemporaneously reported) comprised four cases arriving from Nepal in September 2020: C1, C2, C3, & C4 of which three (C1 – C3) were also linked as family contacts and arrived on the same day while unlinked case C4 arrived four days later. All four cases belonged to lineage B.1.36.27. Cases C2, C3 & C4 shared 100% pair-wise sequence identity, with case C1 two genome-wise substitutions from and phylogenetically basal to the three identical cases implicating C1 as the probable source of the cluster. This is also supported by the relative epidemiological overlap shown in Fig. 2. Notably, case C4 completed quarantine after testing negative on day 13 of 14 and was subsequently linked as the source of infection for three local relatives which likely initiated the fourth epidemic wave of COVID-19 in Hong Kong.^16^

**Figure 2.**
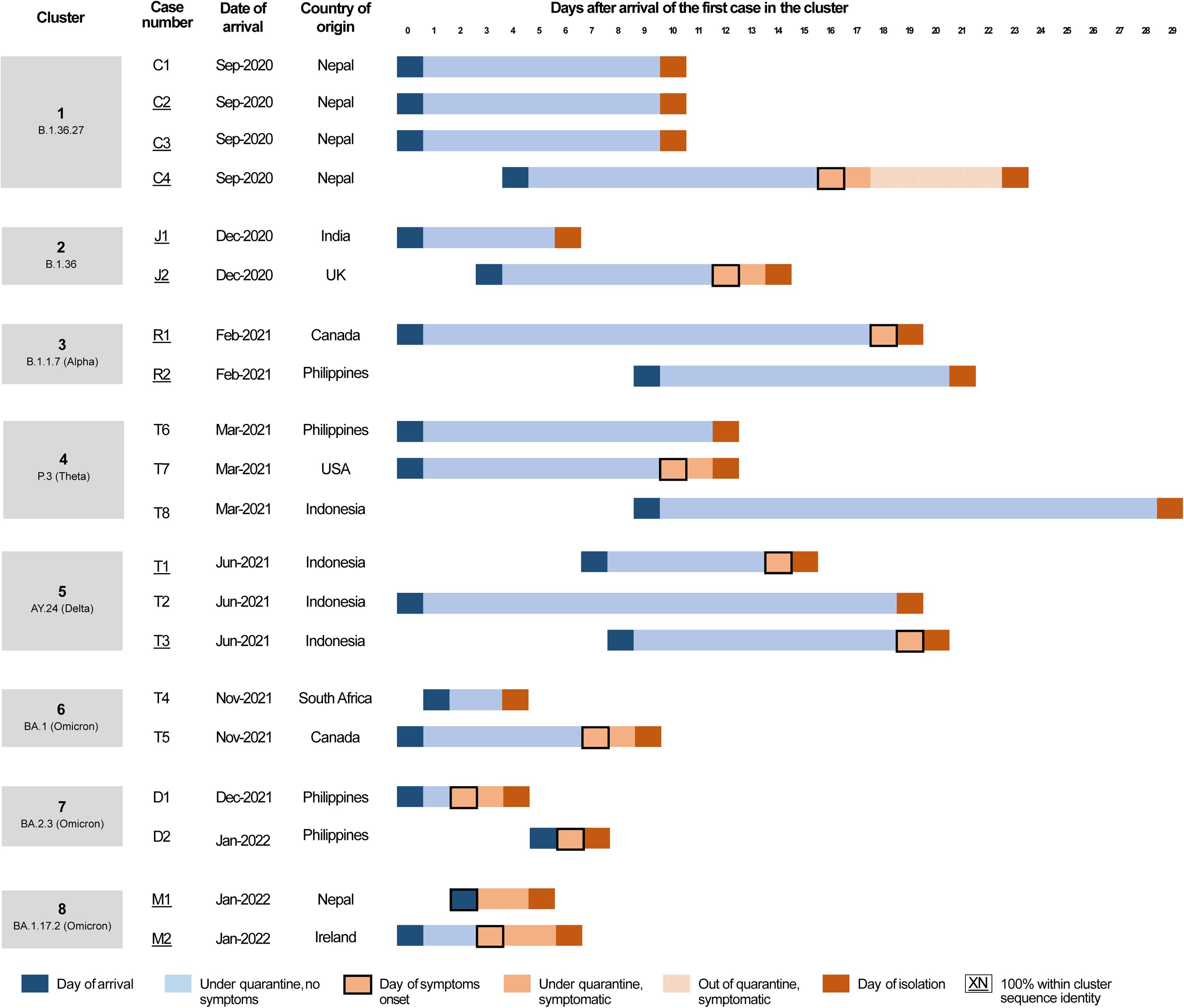
Cases of confirmed SARS-CoV-2 transmission between epidemiologically unlinked arrivals within hotel quarantine (HQ) or designated hotel quarantine (DHQ) in Hong Kong from 1 May 2020 through 31 Jan 2022. Case number in each cluster was ordered by date of reporting (Supplementary Table 2).

Cluster two included cases J1 & J2. Case J1 arrived in Hong Kong from India in December 2020 and case J2 arrived from the UK three days later. Despite different countries of departure, both cases had 100% pair-wise sequence identity and belonged to lineage B.1.36. Due to the identical nature of the sequences the direction of transmission could not be determined, however, the relative overlap and from arrival-to-confirmation or onset between the two cases suggest that case J1 was the likely the infector of case J2 (Fig. 2).

Cluster three (contemporaneously reported) involved case R1 & R2. Case R1 arrived from Canada in February 2021, while case R2 arrived from the Philippines nine days later and both were required to undergo 21 days mandatory quarantine due to a policy change (effective from 2 February 2021). As with cluster two, despite different countries of departure, both cases shared 100% pair-wise sequence identity and belonged to lineage B.1.1.7 (Alpha VOC). Given the epidemiological overlap (Fig. 2) and the latent period distribution of SARS-CoV-2 (Median: 5.5 days, upper 95^th^ percentile: 10.6 days),^17^ it is most likely that R2 infected R1.

Cluster four involved three cases T6, T7 and T8. Cases T6 and T7 arrived in Hong Kong on the same day in March 2021, from the Philippines and the USA respectively, and were required to undergo 21 days mandatory quarantine. Both sequences belonged to lineage P.3 (Theta VOC) despite the different countries of departure and were separated by only two genome-wise substitutions (Fig.2). The third case arrived from Indonesia nine days after T6 and T7, and the sequence was separated by two or three substitutions between T6 and T7 respectively suggesting that T6 was the likely source of infection of T7 and T8. Case T8 was asymptomatic throughout their quarantine period.

Cluster five comprised two or three cases: T1, T2, and T3. These arrived from Indonesia with T2 arriving first, T1 arriving seven days later, and T3 arriving eight days later. T1 and T3 shared 100% sequence identity and were thus considered probable HQ/DHQ transmission though both arrived separately from Indonesia, while T2 was separated by two substitutions so within HQ/DHQ transmission was considered less likely. All three belonged to lineage AY.24, a Delta-like (B.1.617.2) subvariant. Given the dates of arrival and symptom onset, it is possible case T1 was the infector of case T3, though if transmission with the third case is considered, T2 may be the infector of both T1 and T3 (Fig.1, Fig.2).

The last three clusters (six to eight) each comprised two sequenced cases and belonged to Omicron variant lineages or sub lineages of BA.1 and BA.2. Cluster six (contemporaneously reported) an early BA.1 Omicron comprised two cases T4 and T5 arriving from South Africa and Canada in November 2021 and differed by only one nucleotide substitution.^18^ Evidence for cluster seven (BA.2.3) was weaker, with both cases, D1 and D2, arriving from the Philippines separately. Sequences differed by two substitutions though cases had no epidemiological overlap during their stay but was instead grouped into a large cluster with 22 other cases missing sequence data. This suggested perhaps an unsequenced case may have infected both D1 and D2 or less likely acted as an intermediate. The eighth and final cluster (BA.1.17.2) comprised two cases, M1 and M2, who arrived separately from Nepal and Ireland in January 2022. Virus sequences were genomically identical consistent with within-DHQ transmission.

### Variants of concern

Overall, among the 221 sequenced cases with suspected HQ/DHQ overlap to other unlinked cases, 158 were positive for a VOC, the most common being Delta (B.1.617.2-like, n=42/158, 27%) and Alpha (B.1.1.7-like, n = 38/158, 24%). There was no evidence that sequenced HQ/DHQ transmission clusters were more likely to involve a VOC compared to imported cases detected in HQ/DHQ (OR = 0.92, 95% CI = 0.35 2.72, p = 0.87, Logistic regression, Supplementary Table 1).

## DISCUSSION

In this study, we present the descriptive epidemiology of all COVID-19 cases imported into Hong Kong between 1 May 2020 and 31 January 2022 and identify instances of probable SARS-CoV-2 transmission between persons staying within HQ/DHQ. Most significantly, among the 221 imported cases with available genomic data across 49 potential transmission clusters given the time and hotel of stay, 20 (9%) were phylogenetically linked into eight probable clusters, five of which were previously unrecognised. Beyond the eight clusters reported in our study, six additional instances of likely HQ/DHQ transmission between unlinked guests have also been reported by the Centre for Health Protection in Hong Kong both within and after our study period, ^2,19-22^ bringing the total number of probable HQ/DHQ transmissions in Hong Kong to a minimum of 14. In some of the additional instances, the presumptive HQ/DHQ infectee was confirmed after they had completed mandatory quarantine during periods of limited to no local transmission, thus making local infection post-quarantine highly unlikely.

Most notably, a HQ/DHQ cluster reported in this study (Cluster 1) was responsible for the introduction of B.1.36.27 which initiated the fourth epidemic wave in Hong Kong,^16^ while the fifth wave was initiated by a contemporaneously-recognised DHQ transmission of BA.2^20^ which initiated the fifth and largest wave of COVID-19 in Hong Kong with more than 1.1 million cases and more than 9000 deaths. ^5^ For two of the earlier recognised HQ/DHQ clusters, subsequent environmental investigations raised concerns about ventilation systems and airflow between rooms.^2^ Additionally, there have been instances of staff infected whilst working within HQ/DHQ,^7,23^ and other apparent introductions of novel lineages and variants without identified sources despite strict quarantine measures, including the introductions which resulted in the third epidemic wave in Hong Kong. ^3^ Overall, inbound hotel quarantine was able to prevent a large number of introductions, given that more than a thousand cases were identified in HQ/DHQ with substantially reduced daily arrivals compared to the period before the pandemic, but could not entirely prevent the introduction of Omicron, nor other lineages prior.

At the time of this study, Hong Kong is one of very few jurisdictions continuing to pursue a local elimination strategy termed ‘dynamic zero covid’ in 2022 despite the widespread availability and high uptake of vaccines since early 2021. Prior to the emergence of the Omicron variant, local outbreaks were controlled using a variety of stringent public health and social measures, though in each instance, did not involve the sustained circulation of any VOCs which now dominate global circulation (some local VOC infections were reported during periods of elimination, though in each case were contained using existing measures).^24^ Given the widespread global distribution of highly infectious VOCs with shorter incubation periods^25,26^ and the common experience of other formerly elimination-focused countries,^27^ the success of an ongoing elimination strategy in Hong Kong would be aided by substantial improvements to HQ/DHQ infection control or the construction of purpose-built quarantine facilities. However, at this time, the public health impact of future introductions is likely to be substantially lower given the high rate of vaccine uptake as well as natural immunity resulting from the fifth wave. Whereas earlier in the pandemic, inbound quarantine could be reasonably justified as protecting public health from direct and indirect COVID-19 associated mortality via reducing the introduction of infections into the community and thereby prolonging the time between community epidemics, the instances of within HQ/DHQ infection including those identified in this study demonstrate that Hong Kong remains at non-zero risk of future introductions. The need therefore for ongoing inbound quarantine must now be balanced with the ongoing economic and social impacts of continuing such policies, with their incremental and increasingly limited public health benefits.

Our study has a few limitations. Firstly, valid samples or full-length sequence data was only available for a minority of cases identified as potential episodes of HQ/DHQ transmission (38.9%, n=280/720). Furthermore, among the 95 potential HQ/DHQ clusters identified that had available sequence data, 49 of those clusters had only one sample or comprised only samples of known family contacts and were effectively excluded from any inference. Although many of the excluded cases generally had high Ct-values typically associated with a lower risk of infectivity (personal communication, Ben Cowling), it is possible we may have missed more instances of within HQ/DHQ transmission. If the risk of transmission was equivalent in the available and unavailable or excluded potential HQ/DHQ transmission events, we estimate we might have identified 16 additional (24 total) within HQ/DHQ transmission clusters during the equivalent study period. Furthermore, there were a number of HQ/DHQ cases with abnormally long incubation periods that were missing sequence data. Some of these cases might have resulted from within HQ/DHQ transmission, given the short latent period distribution of SARS-CoV-2 (Median: 5.5 days, upper 95^th^ percentile: 10.6 days).^17^

Second, data on potential contact or proximity of unlinked HQ/DHQ cases such as room or floor of stay within HQ/DHQ was not readily available for all cases, though close proximity of rooms were reported among some clusters. Other analyses outside the study period in Hong Kong have used such data to infer likely HQ/DHQ transmissions in the event of post-HQ/DHQ detection of SARS-CoV-2 cases during periods of elimination.^2^ In our study, such data could have otherwise provided additional epidemiological evidence indicative of within HQ/DHQ infection in the absence of genomic data. Similarly, there was no data on potential contact between unlinked cases prior to arrival in Hong Kong (e.g. HQ/DHQ cluster one arriving from the same country), or within the airport (e.g. HQ/DHQ cluster four arriving on the same day), representing potential alternative sources of infection which would overestimate the number of within HQ/DHQ transmission. However, we did note and exclude clusters with the same date and country of arrival or with identical flight numbers when available. ^24,28^ Future studies and surveillance practice could look to sample all SARS-CoV-2 cases detected in HQ/DHQ and combine both additional sources of epidemiological and genomic data to calculate to underling risk of infection, while quantifying the relative effects of potential risk-mitigation factors such as ventilation, room spacing, and HQ/DHQ capacity.

In conclusion, we report at least eight instances of suspected within HQ/DHQ transmission in Hong Kong, demonstrating the risks associated with confining travellers to HQ/DHQ. Infection prevention and control measures within HQ/DHQ should be reviewed for potential improvements, although the ongoing need for inbound quarantine in mid-2022 should be weighed against its economic and social impacts given increasing population immunity from vaccinations as well as the large number of domestic infections in 2022 to date. In the event of a future pandemic, governments aiming to prevent, or at least delay, local outbreaks of infection could consider the use of purpose-built facilities.

## Data Availability

All data produced in the present study are available upon reasonable request to the authors

## ACKNOWLEDGMENTS

We gratefully acknowledge the staff from the originating laboratories responsible for obtaining the specimens. We acknowledge the technical support provided by colleagues from the Centre for PanorOmic Sciences of the University of Hong Kong. We also acknowledge the Centre for Health Protection of the Department of Health for providing epidemiological data for the study.

## FUNDING

This project was supported by the Health and Medical Research Fund, Food and Health Bureau, Government of the Hong Kong Special Administrative Region (grant no. COVID190205 & COVID190118). The funding bodies had no role in the design of the study, the collection, analysis, and interpretation of data, or writing of the manuscript.

## AUTHOR CONTRIBUTIONS

All authors meet the ICMJE criteria for authorship. The study was conceived by LLMP and BJC. HG analysed the NGS data. DCA wrote the first draft of the manuscript and conducted the combined epidemical and genomic analyses. MMS compiled the epidemiological dataset and assisted in writing of the first draft. All authors provided critical review and revision of the text and approved the final version.

## DATA SHARING STATEMENT

All final genomic sequences are made available online upon publication via GISAID with accession numbers listed in the supplementary material. All individual demographic data of sequenced cases also used in this study after de-identification are available upon publication in Supplementary Table 2.

## DECLARATION OF INTERESTS

BJC consults for AstraZeneca, Fosun Pharma, GlaxoSmithKline, Moderna, Pfizer, Roche and Sanofi Pasteur. The authors report no other potential conflicts of interest.

## Notes

### Author Declarations

IRB approval was obtained from the Institutional Review Board of the University of Hong Kong

