## Appendix for "Within-hotel transmission of SARS-CoV-2 during on-arrival quarantine in Hong Kong"

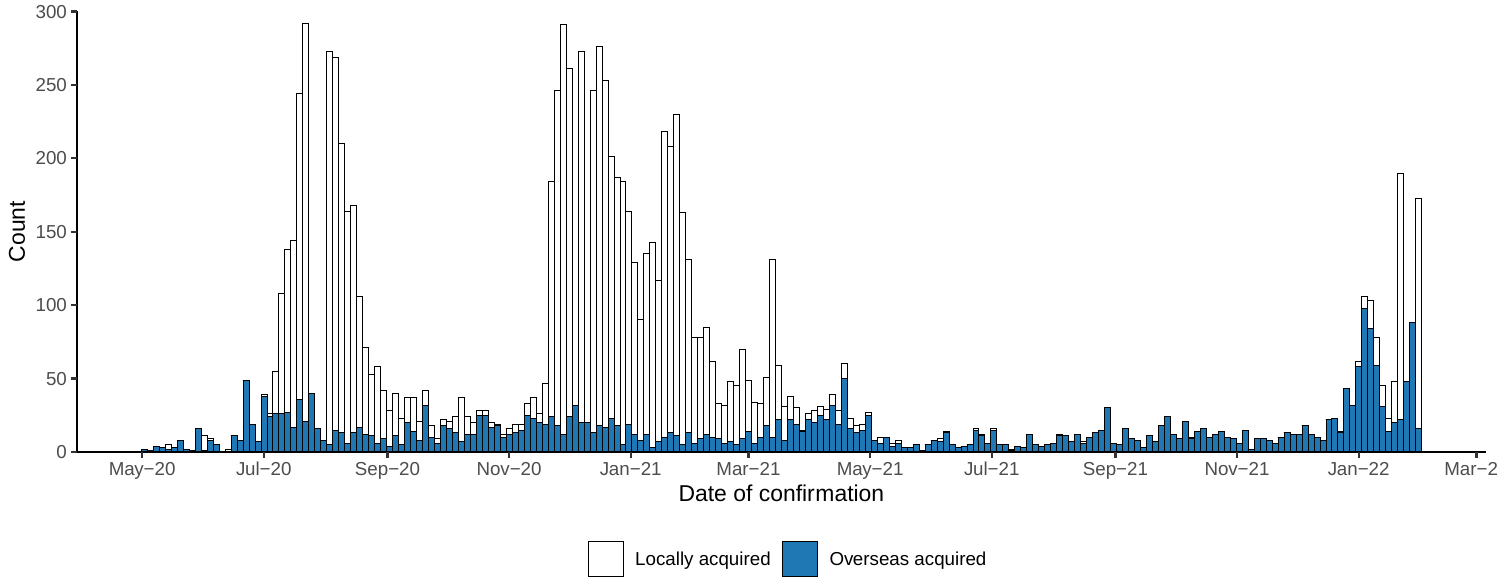

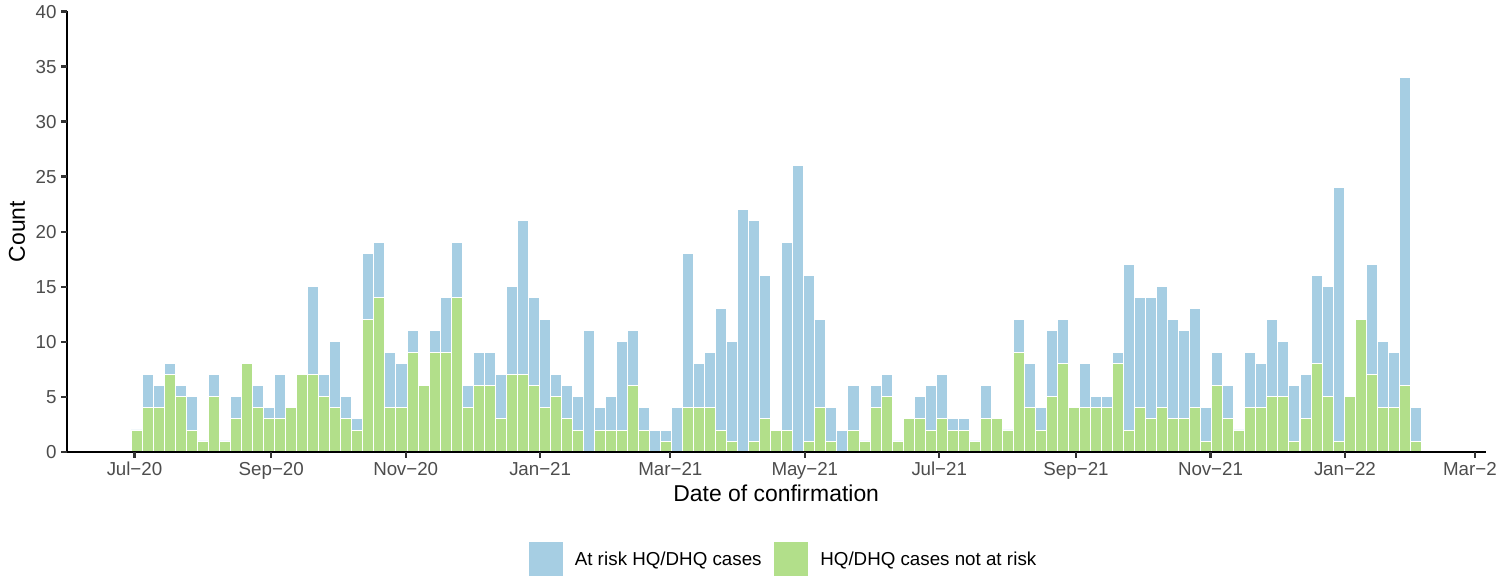

**Supplementary Figure 1. (A)** Epidemic curve of all cases of COVID-19 confirmed within Hong Kong during the study period (1 May 2020 – 31 Jan 2022) by classified source of infection subset. (**B**) Epidemic curve of all cases classified as overseas acquired subset by into DHQ and Non DHQ cases (positive on arrival or not in DHQ) and those at risk of infection within DHQ.

**Supplementary Table 1**. SARS-CoV-2 variants of concern (VOC) identified among cases identified within DHQ transmission clusters and non-DHQ cluster cases. The odds that a DHQ case was phylogenetically linked to a DHQ cluster involving a VOC was 0.92 (95% CI = 0.35, 2.72) though this was not statistically significant (p = 0.87, Logistic regression)

|  | DHQ  cluster cases | non-DHQ  cluster cases | Row  Total |
| --- | --- | --- | --- |
| VOC | 14 | 144 | 158 |
| non-VOC | 6 | 57 | 63 |
| Column Total | 20 | 201 | 221 |

**Supplementary Table 2**. Travel related SARS-CoV-2 cases detected in designated hotel quarantine in Hong Kong between 1 May 2020 and 31 Jan 2022 with available sequence data linked into at risk epidemiological clusters based on hotel of stay and dates or arrival and admission

| Phylo. Cluster | Risk. Cluster | Case | Lineage | Variant | Departure | Hotel | Arrival | Arrival-to-onset | Arrival-to-isolation | Arrival-to-report |
| --- | --- | --- | --- | --- | --- | --- | --- | --- | --- | --- |
| 1 | 1 | C1 | B.1.36.27 |  | Nepal | C | Sep 2020 |  | 10 | 11 |
| 1 | 1 | C2 | B.1.36.27 |  | Nepal | C | Sep 2020 |  | 10 | 11 |
| 1 | 1 | C3 | B.1.36.27 |  | Nepal | C | Sep 2020 |  | 10 | 11 |
| 1 | 1 | C4 | B.1.36.27 |  | Nepal | C | Sep 2020 | 12 | 19 | 21 |
|  | 1 | C5 | B.1.560 |  | India | C | Sep 2020 |  | 10 | 11 |
|  | 1 | C6 | B.1.560 |  | India | C | Sep 2020 |  | 10 | 12 |
| 2 | 2 | J1 | B.1.36 |  | India | J | Dec 2020 |  | 6 | 8 |
| 2 | 2 | J2 | B.1.36 |  | United Kingdom | J | Dec 2020 | 9 | 11 | 12 |
|  | 2 | J3 | B.1.2 |  | USA | J | Dec 2020 | 1 | 6 | 7 |
|  | 2 | J4 | B.1.36.19 |  | Indonesia | J | Jan 2021 | 11 | 14 | 14 |
| 3 | 3 | R1 | B.1.1.7 | Alpha | Canada | R | Feb 2021 | 18 | 19 | 20 |
| 3 | 3 | R2 | B.1.1.7 | Alpha | Philippines | R | Feb 2021 |  | 12 | 13 |
|  | 3 | R5 | B.1.459 |  | Indonesia | R | Feb 2021 |  | 12 | 13 |
|  | 3 | R6 | B.1.466.2 |  | Indonesia | R | Feb 2021 |  | 12 | 13 |
| 4 | 4 | T6 | P.3 | Theta | Philippines | T | Mar 2021 |  | 12 | 13 |
| 4 | 4 | T7 | P.3 | Theta | USA | T | Mar 2021 | 10 | 12 | 13 |
| 4 | 4 | T8 | P.3 | Theta | Indonesia | T | Mar 2021 |  | 19 | 20 |
|  | 4 | T10 | B.1.351 | Beta | Philippines | T | Mar 2021 | 11 | 12 | 13 |
|  | 4 | T11 | B.1.617.1 | B.1.617.1-like | India | T | Apr 2021 |  | 13 | 14 |
|  | 4 | T12 | B.1.617.1 | B.1.617.1-like | India | T | Apr 2021 |  | 13 | 14 |
|  | 4 | T13 | B.1.617.1 | B.1.617.1-like | India | T | Apr 2021 |  | 14 | 14 |
|  | 4 | T14 | B.1.1.7 | Alpha | Pakistan | T | Apr 2021 |  | 13 | 13 |
|  | 4 | T15 | B.1.1.7 | Alpha | USA | T | Apr 2021 | 10 | 12 | 13 |
|  | 4 | T9 | P.3 | Theta | Philippines | T | Mar 2021 |  | 12 | 13 |
| 5 | 5 | T1 | AY.24 | Delta | Indonesia | T | Jun 2021 | 7 | 8 | 9 |
| 5 | 5 | T2 | AY.24 | Delta | Indonesia | T | Jun 2021 |  | 19 | 20 |
| 5 | 5 | T3 | AY.24 | Delta | Indonesia | T | Jun 2021 | 11 | 12 | 13 |
|  | 5 | T16 | AY.9 | Delta | United Kingdom | T | Jun 2021 | 3 | 3 | 4 |
| 6 | 6 | T4 | BA.1 | Omicron | South Africa | T | Nov 2021 |  | 3 | 4 |
| 6 | 6 | T5 | BA.1 | Omicron | Canada | T | Nov 2021 | 7 | 9 | 10 |
| 7 | 7 | D1 | BA.2.3 | Omicron | Philippines | D | Dec 2021 | 2 | 4 | 4 |
| 7 | 7 | D2 | BA.2.3 | Omicron | Philippines | D | Jan 2022 | 1 | 2 | 3 |
| 8 | 8 | M1 | BA.1.17.2 | Omicron | Nepal | M | Jan 2022 | 0 | 3 | 4 |
| 8 | 8 | M2 | BA.1.17.2 | Omicron | Ireland | M | Jan 2022 | 3 | 6 | 8 |
|  | 9 | B1 | P.3 | Theta | Philippines | B | Mar 2021 | 5 | 12 | 13 |
|  | 9 | B2 | B.1.1.7 | Alpha | Philippines | B | Mar 2021 | 11 | 13 | 13 |
|  | 9 | B3 | B.1.351 | Beta | Philippines | B | Mar 2021 |  | 13 | 13 |
|  | 10 | E1 | B.1.36 |  | Pakistan | E | Nov 2020 |  | 17 | 18 |
|  | 10 | E2 | B.1.36 |  | India | E | Dec 2020 |  | 13 | 14 |
|  | 10 | E3 | B.1.36 |  | India | E | Dec 2020 |  | 13 | 14 |
|  | 11 | G1 | B.1.1.63 |  | Philippines | G | Jul 2020 |  | 1 | 2 |
|  | 11 | G2 | B.1.1.63 |  | Philippines | G | Jul 2020 |  | 1 | 2 |
|  | 11 | G3 | B.1.1.263 |  | Philippines | G | Jul 2020 | 1 | 1 | 2 |
|  | 12 | G4 | B.1.36 |  | India | G | Jul 2020 |  | 2 | 1 |
|  | 12 | G5 | B.1.1.306 |  | India | G | Jul 2020 |  | 5 | 2 |
|  | 12 | G6 | B.1.1 |  | India | G | Jul 2020 |  | 5 | 1 |
|  | 13 | G7 | B.1.280 |  | USA | G | Dec 2020 | 9 | 12 | 13 |
|  | 13 | G8 | B.1.280 |  | USA | G | Dec 2020 | 4 | 12 | 13 |
|  | 13 | G9 | B.1.459 |  | Indonesia | G | Dec 2020 |  | 12 | 13 |
|  | 14 | H1 | B.1.1.63 |  | Philippines | H | Sep 2020 |  | 10 | 12 |
|  | 14 | H2 | B.1.1.63 |  | Philippines | H | Sep 2020 |  | 10 | 12 |
|  | 15 | H4 | B.1.36 |  | India | H | Dec 2020 | 11 | 12 | 13 |
|  | 15 | H5 | B.1.1.7 | Alpha | United Kingdom | H | Dec 2020 | 5 | 12 | 13 |
|  | 16 | H6 | B.1.567 |  | USA | H | Dec 2020 |  | 12 | 13 |
|  | 16 | H7 | B.1.567 |  | USA | H | Dec 2020 | 7 | 12 | 13 |
|  | 16 | H8 | B.1.36.29 |  | UAE | H | Jan 2021 | 6 | 14 | 14 |
|  | 17 | J5 | B.1.1.7 | Alpha | Philippines | J | Mar 2021 |  | 13 | 14 |
|  | 17 | J6 | P.3 | Theta | Philippines | J | Mar 2021 |  | 13 | 13 |
|  | 18 | M3 | B.1.1.7 | Alpha | United Kingdom | M | Dec 2020 | 9 | 10 | 12 |
|  | 18 | M4 | B.1.36 |  | Nepal | M | Dec 2020 |  | 13 | 13 |
|  | 18 | M5 | B.1.1.214 |  | Japan | M | Dec 2020 | 1 | 12 | 13 |
|  | 19 | M10 | B.1.1.7 | Alpha | Philippines | M | Mar 2021 |  | 12 | 13 |
|  | 19 | M11 | B.1.351 | Beta | Philippines | M | Mar 2021 |  | 12 | 13 |
|  | 19 | M6 | B.1.562 |  | India | M | Feb 2021 |  | 13 | 14 |
|  | 19 | M7 | B.1.351 | Beta | Philippines | M | Feb 2021 |  | 12 | 13 |
|  | 19 | M8 | B.1.1.7 | Alpha | UAE | M | Mar 2021 | 3 | 3 | 5 |
|  | 19 | M9 | B.1 |  | Indonesia | M | Mar 2021 | 11 | 12 | 13 |
|  | 20 | Q1 | B.1 |  | Pakistan | Q | Aug 2020 |  | 15 | 16 |
|  | 20 | Q2 | B.1.1.63 |  | Philippines | Q | Aug 2020 | 8 | 10 | 11 |
|  | 20 | Q3 | B.1.1.63 |  | Philippines | Q | Aug 2020 | 10 | 10 | 11 |
|  | 21 | Q4 | B.1.1.216 |  | Nepal | Q | Oct 2020 | 6 | 10 | 12 |
|  | 21 | Q5 | B.1 |  | Pakistan | Q | Oct 2020 |  | 10 | 11 |
|  | 22 | Q10 | B.1.617.1 | B.1.617.1-like | Indonesia | Q | Mar 2021 | 19 | 20 | 20 |
|  | 22 | Q6 | B.1 |  | Indonesia | Q | Mar 2021 |  | 1 | 2 |
|  | 22 | Q7 | B.1.1.7 | Alpha | Philippines | Q | Mar 2021 |  | 13 | 13 |
|  | 22 | Q8 | B.1.1.7 | Alpha | Philippines | Q | Mar 2021 |  | 13 | 13 |
|  | 22 | Q9 | B.1.1.7 | Alpha | Philippines | Q | Mar 2021 |  | 13 | 14 |
|  | 23 | R3 | B.1.36 |  | India | R | Sep 2020 |  | 10 | 11 |
|  | 23 | R4 | B.1.36 |  | India | R | Sep 2020 |  | 10 | 11 |
|  | 24 | R10 | B.1.1.7 | Alpha | India | R | Apr 2021 |  | 5 | 5 |
|  | 24 | R11 | B.1.617.1 | B.1.617.1-like | India | R | Apr 2021 | 6 | 7 | 7 |
|  | 24 | R12 | B.1.617.1 | B.1.617.1-like | India | R | Apr 2021 | 5 | 5 | 7 |
|  | 24 | R13 | B.1.617.1 | B.1.617.1-like | India | R | Apr 2021 |  | 6 | 8 |
|  | 24 | R14 | B.1.617.1 | B.1.617.1-like | India | R | Apr 2021 |  | 7 | 8 |
|  | 24 | R15 | B.1.617.1 | B.1.617.1-like | India | R | Apr 2021 |  | 6 | 8 |
|  | 24 | R16 | B.1.1.7 | Alpha | Philippines | R | Mar 2021 |  | 12 | 13 |
|  | 24 | R17 | B.1.617.1 | B.1.617.1-like | India | R | Apr 2021 |  | 11 | 11 |
|  | 24 | R18 | B.1.617.2 | Delta | India | R | Apr 2021 |  | 12 | 13 |
|  | 24 | R19 | B.1.617.1 | B.1.617.1-like | India | R | Apr 2021 |  | 13 | 14 |
|  | 24 | R20 | B.1.617.1 | B.1.617.1-like | India | R | Apr 2021 |  | 13 | 14 |
|  | 24 | R21 | B.1.617.1 | B.1.617.1-like | India | R | Apr 2021 |  | 13 | 14 |
|  | 24 | R22 | B.1.617.1 | B.1.617.1-like | India | R | Apr 2021 |  | 13 | 14 |
|  | 24 | R23 | B.1.617.1 | B.1.617.1-like | India | R | Apr 2021 |  | 13 | 14 |
|  | 24 | R24 | B.1.617.1 | B.1.617.1-like | India | R | Apr 2021 |  | 13 | 14 |
|  | 24 | R25 | B.1.617.1 | B.1.617.1-like | India | R | Apr 2021 |  | 13 | 14 |
|  | 24 | R26 | B.1.617.1 | B.1.617.1-like | India | R | Apr 2021 |  | 14 | 16 |
|  | 24 | R27 | B.1.617.2 | Delta | India | R | Apr 2021 |  | 12 | 13 |
|  | 24 | R7 | B.1.1.7 | Alpha | India | R | Mar 2021 |  | 12 | 13 |
|  | 24 | R8 | B.1.1.7 | Alpha | India | R | Mar 2021 |  | 12 | 13 |
|  | 24 | R9 | B.1.351 | Beta | Philippines | R | Mar 2021 |  | 12 | 14 |
|  | 25 | S1 | B.1 |  | USA | S | Oct 2020 | 9 | 10 | 11 |
|  | 25 | S2 | B.1 |  | Pakistan | S | Oct 2020 | 8 | 10 | 11 |
|  | 26 | S3 | B.1.1.7 | Alpha | United Kingdom | S | Dec 2020 | 3 | 3 | 5 |
|  | 26 | S4 | B.1.1.7 | Alpha | UK | S | Dec 2020 | 6 | 8 | 9 |
|  | 26 | S5 | B.1.1.7 | Alpha | United Kingdom | S | Dec 2020 |  | 8 | 9 |
|  | 26 | S6 | B.1.1.216 |  | India | S | Dec 2020 |  | 12 | 13 |
|  | 27 | S7 | B.1.1.12 |  | Indonesia | S | Jan 2021 | 3 | 14 | 13 |
|  | 27 | S8 | B.1.1.63 |  | Philippines | S | Jan 2021 |  | 12 | 13 |
|  | 28 | S10 | B.1.1.7 | Alpha | Philippines | S | Mar 2021 | 7 | 12 | 13 |
|  | 28 | S11 | B.1.1.7 | Alpha | Philippines | S | Mar 2021 |  | 13 | 14 |
|  | 28 | S12 | B.1.1.7 | Alpha | Philippines | S | Mar 2021 | 10 | 12 | 13 |
|  | 28 | S13 | B.1 |  | Indonesia | S | Mar 2021 |  | 12 | 13 |
|  | 28 | S14 | B.1.1.7 | Alpha | Philippines | S | Mar 2021 |  | 13 | 14 |
|  | 28 | S15 | B.1.1.7 | Alpha | India | S | Mar 2021 |  | 12 | 13 |
|  | 28 | S16 | B.1.351 | Beta | Philippines | S | Mar 2021 |  | 12 | 13 |
|  | 28 | S17 | B.1.617.1 | B.1.617.1-like | India | S | Apr 2021 | 4 | 5 | 6 |
|  | 28 | S18 | B.1.617.1 | B.1.617.1-like | India | S | Apr 2021 | 6 | 6 | 8 |
|  | 28 | S19 | B.1.1.7 | Alpha | Philippines | S | Apr 2021 |  | 12 | 13 |
|  | 28 | S20 | B.1.617.1 | B.1.617.1-like | India | S | Apr 2021 |  | 14 | 14 |
|  | 28 | S21 | B.1.617.1 | B.1.617.1-like | India | S | Apr 2021 |  | 13 | 14 |
|  | 28 | S22 | B.1.617.1 | B.1.617.1-like | India | S | Apr 2021 |  | 14 | 14 |
|  | 28 | S23 | B.1.617.1 | B.1.617.1-like | India | S | Apr 2021 |  | 13 | 14 |
|  | 28 | S24 | B.1.617.1 | B.1.617.1-like | India | S | Apr 2021 |  | 9 | 9 |
|  | 28 | S25 | B.1.617.2 | Delta | Nepal | S | Apr 2021 | 5 | 7 | 9 |
|  | 28 | S26 | B.1.617.2 | Delta | Nepal | S | Apr 2021 | 5 | 8 | 9 |
|  | 28 | S27 | B.1.466.2 |  | Indonesia | S | Apr 2021 |  | 13 | 14 |
|  | 28 | S9 | B.1.525 | Eta | Togo | S | Mar 2021 |  | 13 | 14 |
|  | 29 | W1 | B.1.466.2 |  | Indonesia | W | Mar 2021 |  | 12 | 13 |
|  | 29 | W2 | B.1.466.2 |  | Indonesia | W | Mar 2021 | 11 | 12 | 13 |
|  | 29 | W3 | B.1.1.7 | Alpha | Philippines | W | Apr 2021 | 11 | 13 | 13 |
|  | 29 | W4 | B.1.351 | Beta | Philippines | W | Apr 2021 |  | 13 | 14 |
|  | 30 | X1 | B.1.1.7 | Alpha | Philippines | X | Mar 2021 | 8 | 13 | 13 |
|  | 30 | X2 | B.1.351 | Beta | Philippines | X | Mar 2021 |  | 13 | 13 |
|  | 30 | X3 | B.1.1.7 | Alpha | Philippines | X | Mar 2021 |  | 12 | 13 |
|  | 30 | X4 | B.1.351 | Beta | Philippines | X | Mar 2021 | 7 | 12 | 13 |
|  | 30 | X5 | B.1.1.7 | Alpha | Philippines | X | Mar 2021 |  | 13 | 13 |
|  | 30 | X6 | B.1.617.1 | B.1.617.1-like | India | X | Apr 2021 |  | 21 | 22 |
|  | 31 | Y1 | B.1.1.25 |  | Bangladesh | Y | Nov 2020 |  | 2 | 3 |
|  | 31 | Y2 | B.1.177 |  | United Kingdom | Y | Nov 2020 |  | 12 | 13 |
|  | 31 | Y3 | B.1 |  | USA | Y | Nov 2020 |  | 12 | 13 |
|  | 32 | Z1 | B.1.36 |  | Nepal | Z | Jan 2021 |  | 14 | 14 |
|  | 32 | Z2 | B.1.468 |  | Nepal | Z | Dec 2020 | 18 | 20 | 20 |
|  | 32 | Z3 | B.1.468 |  | Indonesia | Z | Jan 2021 |  | 12 | 13 |
|  | 33 | A1 | B.1.1.7 | Alpha | Pakistan | A | Apr 2021 |  | 12 | 13 |
|  | 33 | A2 | B.1.466.2 |  | Indonesia | A | Apr 2021 | 3 | 13 | 14 |
|  | 34 | F1 | B.1.351 | Beta | Philippines | F | Mar 2021 | 9 | 12 | 13 |
|  | 34 | F2 | AY.75 | Delta | Nepal | F | Apr 2021 | 6 | 8 | 8 |
|  | 35 | L1 | B.1.617.1 | B.1.617.1-like | India | L | Apr 2021 |  | 10 | 10 |
|  | 35 | L2 | B.1.617.1 | B.1.617.1-like | India | L | Apr 2021 |  | 11 | 12 |
|  | 35 | L3 | B.1.617.1 | B.1.617.1-like | India | L | Apr 2021 |  | 11 | 12 |
|  | 35 | L4 | B.1.617.1 | B.1.617.1-like | India | L | Apr 2021 |  | 12 | 14 |
|  | 35 | L5 | B.1.1.7 | Alpha | Japan | L | Apr 2021 |  | 14 | 15 |
|  | 35 | L6 | B.1.617.2 | Delta | India | L | Apr 2021 |  | 12 | 13 |
|  | 36 | M12 | AY.71 | Delta | India | M | Apr 2021 | 6 | 13 | 13 |
|  | 36 | M13 | B.1.351 | Beta | Kenya | M | Apr 2021 |  | 13 | 14 |
|  | 36 | M14 | B.1.617.1 |  | India | M | Apr 2021 |  | 24 | 24 |
|  | 36 | M15 | B.1.351 | Beta | Philippines | M | Apr 2021 | 5 | 8 | 9 |
|  | 36 | M16 | AY.75 | Delta | Nepal | M | Apr 2021 | 7 | 7 | 8 |
|  | 36 | M17 | AY.75 | Delta | Nepal | M | Apr 2021 | 6 | 7 | 9 |
|  | 36 | M18 | AY.75 | Delta | Nepal | M | Apr 2021 |  | 8 | 9 |
|  | 36 | M19 | B.1.36.27 |  | Nepal | M | Apr 2021 |  | 8 | 9 |
|  | 36 | M20 | B.1.617.2 | Delta | Nepal | M | Apr 2021 |  | 8 | 9 |
|  | 36 | M21 | B.1.617.2 | Delta | Nepal | M | Apr 2021 | 4 | 7 | 9 |
|  | 36 | M22 | AY.75 | Delta | Nepal | M | Apr 2021 |  | 8 | 9 |
|  | 36 | M23 | AY.75 | Delta | Nepal | M | Apr 2021 | 4 | 8 | 9 |
|  | 36 | M24 | B.1.1.7 | Alpha | Nepal | M | Apr 2021 |  | 8 | 9 |
|  | 36 | M25 | B.1.1.7 | Alpha | Nepal | M | Apr 2021 | 3 | 8 | 9 |
|  | 36 | M26 | B.1.617.2 | Delta | Nepal | M | Apr 2021 |  | 7 | 8 |
|  | 37 | O1 | BA.1 | Probable Omicron | Ireland | O | Dec 2021 |  | 5 | 6 |
|  | 37 | O2 | BA.1 | Probable Omicron | United Kingdom | O | Dec 2021 | 2 | 2 | 3 |
|  | 37 | O3 | BA.1.1 | Probable Omicron | Australia | O | Dec 2021 |  | 2 | 3 |
|  | 37 | O4 | BA.1.1 | Probable Omicron | Kazakhstan | O | Dec 2021 |  | 3 | 4 |
|  | 37 | O5 | BA.1.17.2 | Probable Omicron | Pakistan | O | Dec 2021 | 4 | 4 | 6 |
|  | 37 | O6 | BA.1.1 | Omicron | USA | O | Dec 2021 |  | 2 | 3 |
|  | 37 | P1 | B.1.617.2 | Delta | Korea | P | Oct 2021 |  | 2 | 4 |
|  | 37 | P10 | B.1.617.2 | Delta | Finland | P | Nov 2021 | 5 | 7 | 8 |
|  | 37 | P11 | AY.33 | Delta | Tanzania | P | Dec 2021 | 2 | 2 | 3 |
|  | 37 | P2 | B.1.617.2 | Delta | Korea | P | Oct 2021 |  | 2 | 4 |
|  | 37 | P3 | AY.38 | Delta | Philippines | P | Oct 2021 |  | 3 | 4 |
|  | 37 | P4 | AY.99 | Delta | Ukraine | P | Oct 2021 |  | 2 | 4 |
|  | 37 | P5 | B.1.617.2 | Delta | Saudi Arabia | P | Oct 2021 | 6 | 7 | 9 |
|  | 37 | P6 | AY.69 | Delta | Korea | P | Nov 2021 | 9 | 10 | 11 |
|  | 37 | P7 | AY.127 | Delta | Singapore | P | Nov 2021 |  | 1 | 2 |
|  | 37 | P8 | AY.46.5 | Delta | Pakistan | P | Nov 2021 | 5 | 7 | 9 |
|  | 37 | P9 | AY.23 | Delta | Pakistan | P | Nov 2021 |  | 10 | 11 |
|  | 38 | Q11 | B.1.1.7 | Alpha | Turkey | Q | Apr 2021 | 3 | 9 | 10 |
|  | 38 | Q12 | B.1.617.2 | Delta | India | Q | Apr 2021 |  | 12 | 13 |
|  | 38 | Q13 | B.1.351 | Beta | Philippines | Q | Apr 2021 |  | 10 | 12 |
|  | 38 | Q14 | B.1.466.2 |  | Indonesia | Q | Apr 2021 |  | 5 | 6 |
|  | 39 | U1 | B.1.351 | Beta | Philippines | U | Mar 2021 | 9 | 12 | 13 |
|  | 39 | U2 | B.1.617.2 | Delta | India | U | Apr 2021 | 10 | 11 | 13 |
|  | 39 | U3 | AY.75 | Delta | India | U | Apr 2021 |  | 13 | 14 |
|  | 39 | U4 | B.1.470 |  | Indonesia | U | Apr 2021 | 10 | 12 | 17 |
|  | 40 | W5 | AY.108 | Delta | Pakistan | W | Oct 2021 |  | 3 | 4 |
|  | 40 | W6 | AY.108 | Delta | Pakistan | W | Oct 2021 |  | 3 | 6 |
|  | 41 | Y4 | B.1.617.2 | Delta | Philippines | Y | Sep 2021 |  | 3 | 4 |
|  | 41 | Y5 | B.1.617.2 | Delta | Philippines | Y | Sep 2021 |  | 3 | 4 |
|  | 41 | Y6 | B.1.617.2 | Delta | Philippines | Y | Sep 2021 |  | 3 | 4 |
|  | 42 | Z4 | B.1.1.7 | Alpha | Pakistan | Z | Mar 2021 |  | 4 | 5 |
|  | 42 | Z5 | B.1.1.7 | Alpha | Pakistan | Z | Mar 2021 |  | 13 | 13 |
|  | 42 | Z6 | B.1.1.7 | Alpha | Pakistan | Z | Mar 2021 |  | 13 | 13 |
|  | 42 | Z7 | B.1.351 | Beta | Pakistan | Z | Mar 2021 | 12 | 13 | 15 |
|  | 42 | Z8 | B.1.351.3 | Beta | Bangladesh | Z | Mar 2021 |  | 12 | 13 |
|  | 43 | G10 | AY.43 | Delta | United Kingdom | G | Dec 2021 | 2 | 3 | 4 |
|  | 43 | G11 | AY.43 | Delta | Czech | G | Nov 2021 |  | 9 | 10 |
|  | 43 | G12 | BA.2.3 | Omicron | Sweden | G | Jan 2022 |  | 3 | 4 |
|  | 44 | I1 | AY.43 | Delta | United Kingdom | I | Nov 2021 |  | 3 | 4 |
|  | 44 | I2 | AY.25 | Delta | USA | I | Nov 2021 |  | 4 | 4 |
|  | 44 | I3 | AY.4.2 | Delta | United Kingdom | I | Nov 2021 | 3 | 5 | 6 |
|  | 45 | I4 | BA.2.12 | Omicron | India | I | Jan 2022 | 2 | 4 | 4 |
|  | 45 | I5 | BA.1 | Omicron | Italy | I | Jan 2022 | 4 | 9 | 10 |
|  | 46 | K1 | BA.1.17.2 | Omicron | Canada | K | Dec 2021 | 3 | 4 | 5 |
|  | 46 | K2 | BA.2 | Omicron | Australia | K | Jan 2022 | 2 | 3 | 4 |
|  | 46 | K3 | BA.1.17 | Omicron | Switzerland | K | Jan 2022 |  | 5 | 6 |
|  | 47 | N1 | BA.1.21 | Omicron | Ghana | N | Dec 2021 |  | 3 | 4 |
|  | 47 | N2 | BA.1.1 | Probable Omicron | Morocco | N | Dec 2021 |  | 5 | 6 |
|  | 48 | V1 | BA.1.17.2 | Omicron | Philippines | V | Jan 2022 |  | 4 | 4 |
|  | 48 | V2 | BA.2.3 | Omicron | Philippines | V | Jan 2022 | 2 | 4 | 4 |
|  | 49 | X7 | BA.1 | Omicron | Nepal | X | Jan 2022 |  | 6 | 6 |
|  | 49 | X8 | BA.2.2 | Omicron | Nepal | X | Jan 2022 | 4 | 5 | 7 |
